# Post-vaccination SARS-CoV-2 neutralizing antibodies in pregnant women receiving biologics for inflammatory bowel disease

**DOI:** 10.1101/2025.03.05.25323433

**Authors:** Donna E. Leet, Jing Jin, Charles S. Craik, Michael G. Kattah, Millie D. Long, Uma Mahadevan

**Affiliations:** Department of Medicine, University of California, San Francisco, CA, USA; Vitalant Research Institute, San Francisco, CA, USA; Department of Pharmaceutical Chemistry, University of California, San Francisco, CA, USA; Department of Medicine, University of North Carolina, Chapel Hill, NC, USA

## Abstract

Inflammatory bowel disease (IBD) treatments and pregnancy can independently modulate immune responses, but the combined effects on SARS-CoV-2 vaccine-induced immunity are poorly understood. This study explores the efficacy of SARS-CoV-2 vaccination and placental antibody transfer among pregnant women with IBD on biologic therapies. This observational study included pregnant women with and without IBD from the PIANO and PREVENT-COVID studies and their neonates. We assessed anti-SARS-CoV-2 neutralizing antibody titers (NT50) in maternal and cord blood post-vaccination using a pseudotype neutralization assay and calculated placental transfer ratios. A total of 32 pregnant women participated, and 27 were exposed to a biologic medication during pregnancy. Neutralizing antibody titers were similar between biologic-treated and non-treated groups, and biologic-exposed women demonstrated robust placental transfer of neutralizing antibodies. Corticosteroid use during pregnancy was significantly associated with reduced placental transfer efficiency, although this effect was not significant in a sensitivity analysis excluding patients treated with immunomodulators. Vaccination timing and previous SARS-CoV-2 infection impacted maternal and cord antibody levels, with higher titers observed in those vaccinated before pregnancy or infected during pregnancy. Overall, our findings suggest that pregnant women with IBD on biologic therapies mount effective SARS-CoV-2 neutralizing antibody responses similar to their non-biologic-exposed counterparts, with efficient placental transfer. These findings reassure the safety and efficacy of SARS-CoV-2 vaccination in this population, though further research is needed to explore the long-term protective effects of transferred antibodies in neonates. Corticosteroid use, immunomodulator use, and vaccination timing may influence antibody dynamics, underscoring the need for tailored clinical management in this vulnerable population.

## Introduction

Effective SARS-CoV-2 vaccination can significantly reduce the risk of COVID infection and mortality in patients with inflammatory bowel disease (IBD) while also improving neonatal outcomes [1]. Understanding the dynamics of vaccine-induced immunity in pregnant women with IBD is crucial, given the immunomodulating effects of both pregnancy and IBD treatments such as biologics. However, limited data exist on the efficacy of SARS-CoV-2 vaccination in pregnant women with IBD receiving immunosuppressive therapy, particularly regarding the placental transfer of antibodies.

While pregnant and non-pregnant women have shown similar antibody responses to SARS-CoV-2 vaccination [2], several studies have illustrated reduced vaccine responses in patients with IBD taking anti-TNF agents or JAK inhibitors [3,4]. Placental transfer of COVID antibodies in healthy pregnant women has been established to varying degrees [5,6], but whether immunosuppressive therapies impact this process is unknown. Finally, most of these studies have quantified SARS-CoV-2 immunoglobulin levels rather than the examining the quality of antibodies denoted by the neutralizing antibody titer, which is of significant biological relevance in patient outcomes [7]. We evaluated the impact of biologic use on (1) anti-SARS-CoV-2 neutralizing antibody titers in post-vaccination maternal and cord blood and (2) placental transfer of anti-SARS-CoV-2 neutralizing antibodies in a cohort of pregnant women with and without IBD.

## Methods

The study was conducted according to Declaration of Helsinki principles and was approved by the Institutional Review Board of the University of California, San Francisco (#10-00831). Written informed consent was received from participants prior to inclusion in the study. The Pregnancy in Inflammatory Bowel Disease and Neonatal Outcomes (PIANO) study is an ongoing prospective observational study that has enrolled 2018 pregnant women with and without IBD at more than 30 US centers beginning in January 2007. Partnership to Report Effectiveness of Vaccination in populations Excluded from iNitial Trials of COVID (PREVENT-COVID) is a prospective, observational, cohort study of patients with IBD in the United States who have received any COVID-19 vaccine that enrolled 3505 participants with IBD from across the US. Pregnant women in the PIANO study, the PREVENT-COVID study, or who were seen within the UCSF Gastroenterology clinic and who had received at least one SARS-CoV-2 vaccination were eligible. Using patient questionnaires and medical records, we collected (1) maternal demographic variables including age, body mass index, parity, gestational age at delivery and smoking, (2) COVID-related variables including type of vaccination, dates of vaccination(s) before and during pregnancy, incidence and timing of COVID infection during pregnancy, and (3) IBD-related variables including diagnosis, disease activity, and IBD medications. Serum was collected from women, umbilical cord, and in some cases, newborns, at the time of delivery. The sample size in the comparison (non-biologic) arm was limited due to challenges in recruiting patients and their neonates after 2022.

Serum samples were assayed for the presence of anti-SARS-CoV-2 neutralizing antibodies at UCSF or at Vitalant Research Institute (San Francisco, California) using a lentivirus-based pseudotype neutralization assay as previously described [8]. Percentage of infectivity was calculated as a percentage of plasma/serum control. Nonlinear regression curves, 50% neutralization titers (NT50) were calculated in GraphPad Prism (v10.2.3). Placental antibody transfer ratios were calculated as the cord blood NT50 divided by maternal NT50.

Data analysis was conducted using GraphPad Prism (v10.2.3). We used descriptive statistics to characterize biologic-exposed and non-exposed populations, with continuous data reported as median and IQR, and descriptive data as numbers and percentages, unless otherwise stated. For comparison of the demographics data between biologic-exposed and non-exposed patients, we used Mann-Whitney U tests for continuous data and Fisher’s exact tests for descriptive data. For the univariate analysis, the Shapiro-Wilk test was used to determine data distribution normality. The data sets were shown to be non-normally distributed. Therefore, the Kruskal-Wallis test was performed to determine significant differences between three or more groups and the Mann-Whitney U test was performed to compare two groups. Spearman’s rank correlation coefficient was calculated to determine the relationship between two sets of non-parametric data. P-values less than 0.05 were considered significant.

## Results

Between May 2021 and November 2022, a total of 96 women were confirmed eligible and recruited for the study, of which 32 patients enrolled, and 29 had IBD. A total of 27 were on a biologic during pregnancy. The majority of biologic-treated women were on an anti-TNF agent (70.4%). Of the two women with IBD not treated with a biologic, one was treated with mesalamine, and one received azathioprine during pregnancy. Three healthy women without IBD were also included in the no biologic cohort. Cord blood was not collected from two patients. None of the participants had significant cardiopulmonary or metabolic comorbidities. For the IBD group, the mothers’ median age at delivery was 34.4 years, and infant median gestational age at delivery was 39.6 weeks. As expected, the breakdown of disease states between the non-biologic and biologic cohorts were statistically disparate, but there were no significant differences in pregnancy-related, SARS-CoV-2-related, and other IBD-related demographic characteristics between biologic-exposed and non-exposed cohorts (Table 1).

**Table 1.** Cohort demographics.

|  | Receiving<br>Biologic | Not Receiving<br>Biologic |  |
| --- | --- | --- | --- |
|  | n = 27 | n = 5 | p-value |
| Maternal age at delivery, years | 34.4 (31.3 - 36.6) | 36.2 (34.9 - 39.0) | 0.11 |
| BMI prior to pregnancy | 22.0 (20.7 - 24.3) | 22.3 (20.8 - 24.6) | 0.95 |
| Smoking history |  |  |  |
| Never | 22 (81.5) | 5 (100) |  |
| Past/present | 5 (18.5) | 0 (0) | 0.56 |
| Disease state |  |  |  |
| Healthy | 0 (0) | 3 (60) |  |
| Crohn's Disease | 17 (63.0) | 0 (0) |  |
| Ulcerative Colitis | 10 (37.0) | 2 (40) | <0.001 |
| Gestational age at delivery, weeks | 39.6 (39 - 40) | 39.0 (35.2 - 40.4) | 0.28 |
| Parity | 1 (1 - 2) | 2 (1 - 2) | 0.66 |
| Type of delivery |  |  |  |
| Vaginal | 22 (81.5) | 3 (60) |  |
| Caesarean | 5 (18.5) | 2 (40) | 0.3 |
| Biologic during pregnancy |  |  |  |
| Anti-TNF | 19 (70.4) |  |  |
| Anti-IL12/23 | 8 (29.6) | n/a | n/a |
| Steroid exposure | 4 (14.8) | 0 (0) | >0.99 |
| Immunomodulator exposure | 4 (14.8) | 1 (20) | >0.99 |
| Vaccinated during pregnancy | 19 (70.4) | 4 (80) | >0.99 |
| COVID infection during pregnancy | 7 (25.9) | 1 (20) | >0.99 |
| Time from last COVID vaccine or infection to antibody testing, days | 141 (54 - 228) | 98 (51.5 - 153.5) | 0.45 |
| Time from last COVID vaccine to antibody testing, days | 156 (56.5 - 370) | 98 (51.5 - 255.5) | 0.76 |
| Vaccine type / Bivalent reception |  |  |  |
| BNT162b2 | 14 (51.9) | 5 (100) |  |
| mRNA-1273 | 13 (48.1) | 0 (0) | 0.06 |
| Received Bivalent | 4 (14.8) | 0 (0) | >0.99 |
| Active disease | 1 (3.7) | 0 (0) | >0.99 |

Table 1 data are reported as median (IQR) or number (%). Mann-Whitney U tests were used for continuous data and Fisher’s exact tests for descriptive data.

There was no difference in maternal nor cord blood anti-SARS-CoV-2 neutralizing antibody titers (NT50) at delivery between biologic-exposed and non-exposed women (Fig. 1A). Maternal and cord blood NT50 also did not vary by anti-TNF exposure as compared to anti-IL12/23 exposure or no biologic (not shown). Univariate analysis of the biologic-exposed subgroup revealed higher maternal NT50, and a trend toward higher cord blood NT50, in women who completed SARS-CoV-2 vaccination prior to conception as opposed to during pregnancy (Fig. 1B). Those who were infected with SARS-CoV-2 during pregnancy had higher maternal and cord blood NT50 (Fig. 1C). There was a positive correlation between time elapsed since final vaccine dose and maternal and cord blood neutralizing antibody titers, with higher NT50 associated with longer latency periods, although this was not statistically significant (p=0.06, not shown).

**Figure 1.**
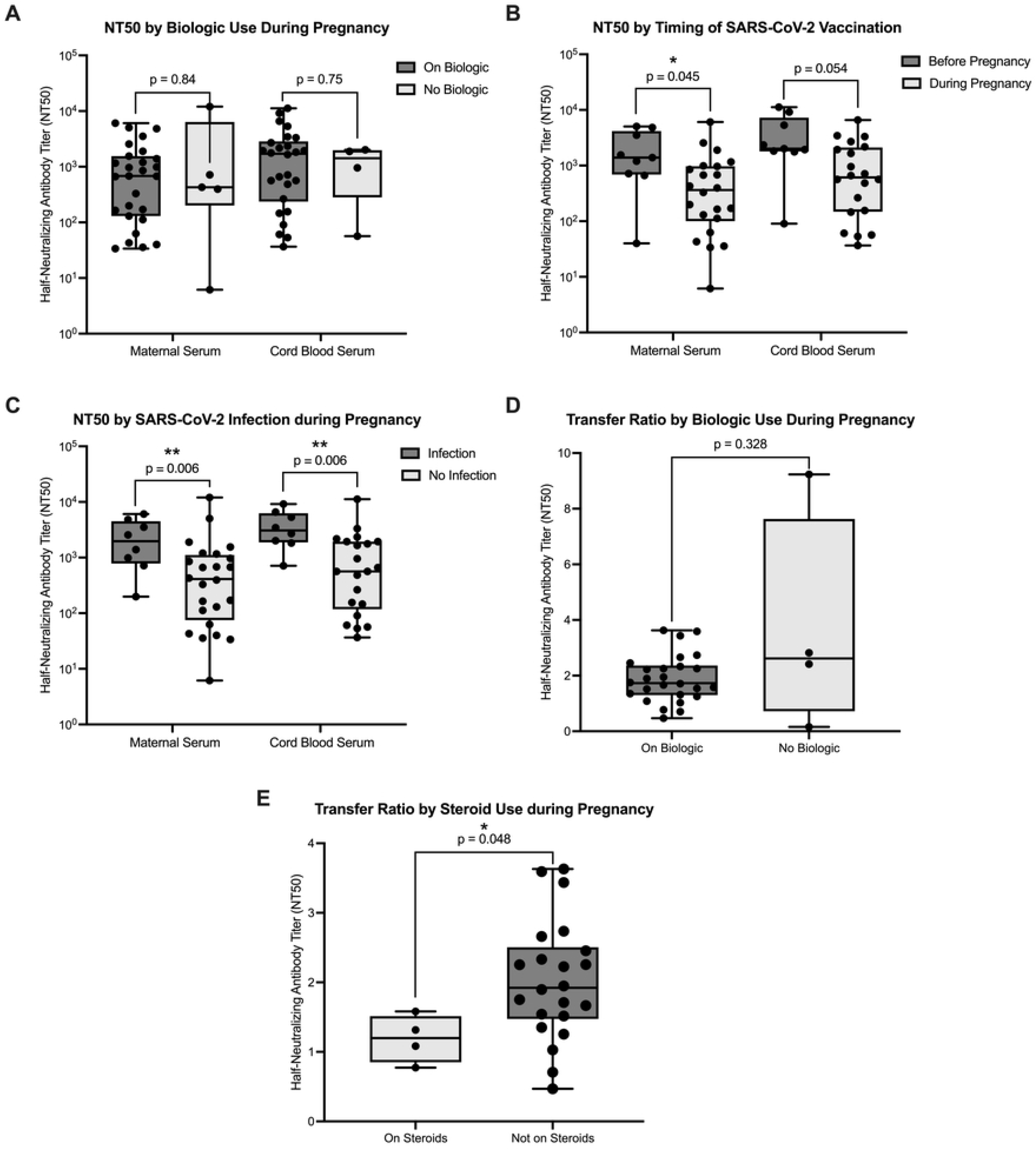
**A**. Maternal and cord blood neutralizing antibody titers (NT50) against SARS-CoV-2 in pregnant women by biologic exposure during pregnancy. **B**. NT50 by timing of SARS-CoV-2 vaccination completion in relation to pregnancy in biologic-exposed mothers. **C**. NT50 by incidence of SARS-CoV-2 infection during pregnancy in biologic-exposed mothers. **D**. Anti-SARS-CoV-2 neutralizing antibody transfer ratio by biologic exposure during pregnancy. **E**. Anti-SARS-CoV-2 neutralizing antibody transfer ratio by steroid use during pregnancy in biologic-exposed mothers. Cord blood was not collected for one patient on biologic and one patient with IBD not on a biologic. Mann-Whitney U tests were performed.

Overall, there was a significant positive correlation between maternal and cord blood neutralizing antibody maternal titers (r = 0.96, p < 0.001, not shown). Placental transfer of neutralizing antibodies occurred in all biologic-treated women. As a measure of transfer efficiency, cord to maternal transfer ratios were calculated. There was no significant difference in transfer ratio based on biologic exposure during pregnancy (Fig. 1D). However, a trend toward a higher transfer rate was noted in the non-biologic arm, although this is likely due to an outlier mother with a low neutralizing antibody titer yet a high neonate titer, leading to a high transfer ratio. In biologic-treated women, the median transfer ratio was greater than 1.5 (1.73). In biologic-exposed women, corticosteroid exposure during pregnancy was significantly associated with a lower placental transfer ratio (Fig. 1E). There was no significant correlation between maternal NT50 or cord blood NT50 and transfer ratio nor between vaccine latency and transfer ratio.

Given the previously illustrated impact of immunomodulators on SARS-CoV-2 antibody titers, we performed a sensitivity analysis excluding immunomodulator-treated patients [3]. We found that the impact of timing of vaccination on maternal NT50 and the effect of steroids on transfer ratio in biologic-exposed women became non-significant (p=0.06 and p=0.17, respectively). Similarly, these effects became non-significant when patients treated with mRNA-1273 vaccine and the BNT162b2 mRNA SARS-CoV-2 vaccine were analyzed separately.

## Discussion

This study highlights important dynamics of neutralizing antibodies in pregnant women with IBD vaccinated against SARS-CoV-2. We found that biologic-exposed pregnant women with IBD were able to mount a neutralizing antibody response similar to their non-biologic exposed counterparts. These results are in line with a prior study of anti-SARS-CoV-2 IgG levels in pregnant women with IBD, of which approximately half were on a biologic [9], but is in contrast to studies of non-pregnant patients with IBD on biologics, which have shown reduced levels of anti-SARS-CoV-2 IgG [3] and neutralizing antibodies [4] in infliximab (anti-TNF)-treated patients. Thus, pregnancy may be attenuating the negative impact of biologic exposure on antiviral immune response in these women.

Our study is the first to show robust placental transfer of anti-SARS-CoV-2 neutralizing antibodies in biologic-treated women, at rates similar to those of women who were vaccinated during pregnancy reported in the literature (ranging 0.77 to 2.6) [5,6,10]. These results also align with established transfer ratios for other vaccinations, such as measles, influenza, and pertussis, which all induce higher cord titers at delivery compared to maternal titers, with favorable (>1) TRs of 1.2–3 [11].

Univariate analysis of our biologic-exposed cohort highlighted that mothers who completed SARS-CoV-2 vaccination before pregnancy had significantly higher neutralizing antibody titers at delivery than those who completed vaccination during pregnancy, with the same trend in cord blood. However, we found no significant correlation between transfer ratio and vaccination timing. While most studies of SARS-CoV-2 IgG titers in women vaccinated during early pregnancy have conversely demonstrated decreasing maternal and neonatal antibody titers with latency from vaccination, antibody transfer ratios have been found to increase with latency from vaccination and infection [6,11]. Importantly, these studies did not examine women who were vaccinated before pregnancy, thus our finding of increased neutralizing antibody titers in women vaccinated prior to pregnancy may reflect a combinatorial immunosuppressive effect of biologic exposure and pregnancy, and/or simply a difference between neutralizing and non-neutralizing antibody dynamics, which has also been illustrated [11,12].

Supporting a potential negative impact of immunosuppressive medications on placental antibody transfer, we found that corticosteroid exposure during pregnancy in biologic-exposed women significantly reduced transfer ratios. However, in our sensitivity analysis in which immunomodulator-treated patients were excluded, this finding became non-significant, suggesting that the combined use of different immunosuppressive agents (corticosteroids and immunomodulators) might have a cumulative impact on placental antibody transfer. While previous research has highlighted impacts of maternal factors such as chronic infection (including HIV, malaria), malnutrition, and metabolic disease on FcRn-mediated transplacental transfer [13], this is the first study to highlight a potential negative impact of corticosteroid and use during pregnancy in biologic and immunomodulator-exposed patients.

Consistent with prior studies of COVID-19 infection in non-vaccinated versus vaccinated pregnant women, we found that vaccinated mothers who had COVID-19 during pregnancy exhibited higher neutralizing antibody titers, demonstrating a boosting effect of infection on antiviral immune response in vaccinated pregnant women [10].

This study faces several limitations that impact the interpretability of its findings. The small sample size, particularly in the non-biologic cohort, restricts generalizability, making it difficult to isolate the effects of biologic versus immunomodulator exposure or vaccine versions. Additionally, longitudinal measurements of neutralizing antibodies in mothers and infants after delivery could inform our understanding of the persistence and efficacy of these antibodies over time. Finally, incorporating newborn COVID-19 outcomes could provide additional context about the impact of maternal antibody transfer on neonatal health.

Overall, biologic treatment during pregnancy in women with IBD does not appear to impair the maternal or neonatal neutralizing antibody response to SARS-CoV-2, but combined steroid and immunomodulator exposure as well as vaccination timing may impact the placental transfer of neutralizing antibodies in this group. Future studies with larger cohorts and longitudinal follow-up are needed to assess if neutralizing antibody titers correlate with improved protection from COVID in biologic-exposed mothers and their neonates.

## Data Availability

All relevant data are within the manuscript and its Supporting Information files.

## Acknowledgements

Jessica Lim, Nicole Arima, George Rutherford, Beatrice Huang, and participating patients.

